# Association of Serum Vitamin D Levels with Depression in Adults: A Nationwide Population-based Study in Korea

**DOI:** 10.1101/2020.10.22.20217968

**Authors:** Young-Eun Jung, Ashley K. Dores, Scott B. Patten, Lakshmi N. Yatham, Raymond W. Lam

## Abstract

**Background:** Vitamin D status may be associated with depression, but there have been inconsistencies in the reported estimates. This study aimed to examine the association of vitamin D status with depression in a large general population sample.

**Methods:** Cross-sectional data for a representative Korean sample of 1,825 people aged 19 years or older were obtained from the nationally representative Korean National Health and Nutrition Examination Survey (2014). Depression was defined by Patient Health Questionnaire-9 (PHQ-9) scores ≥10 (moderate/severe). Logistic regression was used to estimate the associations between depression and serum 25-hydroxyvitamin D [25(OH)D] levels. Continuous serum 25(OH)D level was categorized into quartiles. Continuous PHQ-9 scores were assessed using quantile regression. Adjustments for age, sex, marital status, level of education, lowest income quartile, body mass index, level of physical activity, chronic conditions, serum creatinine level, glomerular filtration rate, and history of depression were used in the statistical analyses. Estimates of prevalence and odds ratios (OR) with 95% confidence intervals (CI) were made. Sampling weights were utilized to account for survey design effects.

**Results:** Individuals in the lowest serum 25(OH)D quartile level had significantly higher prevalence of depression than in the upper three quartiles (8.3% vs. 5.1%; p=0.024). No association was observed between serum 25(OH)D level and depression after adjusting for potential covariates (OR 1.48, 95%CI 0.93, 2.35; p=0.097]. However, a stronger association was observed among male respondents, with an estimated OR of 2.54 (95% CI 1.17, 5.50; p=0.018). Additionally, in the quantile regression analysis, estimates from adjusted models remained significant (β = −0.056, p=0.002).

**Conclusion:** While our findings support the association between lower vitamin D status and depression in Korean adults, additional studies are needed to clarify this relationship.

## 1. Introduction

Major depressive disorder (MDD) is associated with significant disability, mortality, and healthcare costs (Cassano and Fava, 2002; Chang et al., 2012). Although biological, psychological, and environmental theories have been advanced, the underlying pathophysiology of depression has not yet been fully elucidated. Vitamin D is a unique neurosteroid hormone with roles in numerous brain processes, including neuroimmunomodulation, regulation of neurotropic factors, neuroprotection, neuroplasticity, and brain development (Fernandes de Abreu et al., 2009). Receptors for vitamin D are present on neurons and glia in many areas of the brain, including the prefrontal cortex, amygdala, hippocampus, hypothalamus, and cerebellum (Eyles et al., 2005; Zehnder et al., 2001), which have been implicated in the pathophysiology of psychiatric conditions such as depression, schizophrenia, and cognitive dysfunction (Lerner et al., 2018).

Several recently published epidemiological studies have demonstrated an inverse association between serum vitamin D level and depression. Two prospective large cohort studies found that low serum vitamin D levels were associated with the presence and severity of depressive symptoms (Milaneschi et al., 2014; Milaneschi et al., 2010). A number of large cross-sectional studies also have indicated an association between low serum vitamin D level and depressive symptoms (Ganji et al., 2010; Hoang et al., 2011; Hoogendijk et al., 2008; Jaaskelainen et al., 2015; Kjaergaard et al., 2011; Song et al., 2016; Stewart and Hirani, 2010). Moreover, two meta-analyses demonstrated that low vitamin D status is associated with depressive symptoms and increased risk of experiencing a depressive episode (Anglin et al., 2013; Ju et al., 2013). In contrast to these studies reporting significant associations between vitamin D status and depression, other studies have failed to show any such association (Nanri et al., 2009; Pan et al., 2009; Park et al., 2016; Zhao et al., 2010). Further, a recent large prospective cohort study indicated a cross-sectional, but not prospective, association between serum vitamin D level and depressive symptoms (Jovanova et al., 2017).

The inconsistent results from these studies may be related to differences in instruments used to assess depression and the type of assays used to measure vitamin D levels. In addition, vitamin D status is associated with many sociodemographic, lifestyle, and health-related variables. There is also high variation in vitamin D status within countries, geographic regions, and subgroups with specific characteristics, such as ethnicity and skin color (Hilger et al., 2014). Previous epidemiological studies of the associations of vitamin D status with depressive symptoms were more commonly performed in Western populations (Ganji et al., 2010; Hoang et al., 2011; Hoogendijk et al., 2008; Jaaskelainen et al., 2015; Jovanova et al., 2017; Kjaergaard et al., 2011; Milaneschi et al., 2014; Milaneschi et al., 2010; Stewart and Hirani, 2010; Zhao et al., 2010), and focused on older persons (Hoogendijk et al., 2008; Jovanova et al., 2017; Milaneschi et al., 2010; Pan et al., 2009; Song et al., 2016; Stewart and Hirani, 2010).

Studies conducted in East Asian populations (i.e., Japan, China, and Korea) tended to show no associations of vitamin D status with depressive symptoms (Nanri et al., 2009; Pan et al., 2009; Park et al., 2016). However, these studies cannot necessarily be considered representative of the general population (Nanri et al., 2009; Pan et al., 2009) and lacked a relevant standardized measurement of depressive symptoms (Park et al., 2018). This underscores the need for large-scale East Asian studies that include a comprehensive set of potential determinants that may confound the association with vitamin D. Hence, our aim in the present study was to investigate the association between vitamin D status and depression using a validated measure of depressive symptoms and adjusting for potential confounding factors in a representative population sample of Korean adults.

## 2. Methods

### 2.1. Data source and study sample

This study was based on cross-sectional data from the Korean National Health and Nutrition Examination Survey (KNHANES), a nationally representative survey conducted by the Korea Centers for Disease Control and Prevention (KCDC) since 1998. KNHANES was designed to target the non-institutionalized Korean civilian population, and consisted of a health interview survey, a nutrition survey, and a health examination survey. All health interviews and examinations, including blood sampling, were performed in the Mobile Examination Centers on the same day (Kweon et al., 2014).

We used data from the second year of the sixth KNHANES (VI-2, 2014). KNHANES VI-2 used a two-stage stratified cluster sampling procedure. In the first stage, from the 303,180 geographically defined sampling units, 192 primary sampling units (PSUs) were sampled based upon administrative districts and housing types. Each PSU contains an average of 60 households, and with intra-stratification of residential area, age and gender, a systematic sampling system was then applied to extract 20 households from each PSU. In this data set, approximately one-third of the total sample was selected using stratified subsampling for measurement of serum vitamin D levels. We selected for adults aged ≥19 years.

### 2.2. Measurement of depression

The presence of depression was identified using the Patient Health Questionnaire-9 (PHQ-9), which is a reliable and valid assessment tool for measuring depression severity over the preceding 2 weeks (Kroenke et al., 2001), and with items that align with the symptom-based ‘A’ criteria for major depressive episode. The PHQ-9 is composed of nine items rated from 0 (*not at all*) to 3 (*having the symptoms nearly every day*), and the scores for each item are summed to produce a total depression severity score (range: 0–27). In original validation study, a score of 10 or higher had a sensitivity of 88% and a specificity of 88% for detecting major depressive disorder (Kroenke et al., 2001). The optimal cult-off score identified in the Korean validation study was also 10 to achieve a sensitivity of 81.8% and a specificity of 89.9% for detecting major depressive episode (Choi et al., 2007). Participants in this study were classified as having depression if their PHQ-9 score was ≥10.

### 2.3. Measurement of vitamin D status

Serum 25-hydroxyvitamin D [25(OH)D] level is currently considered a reliable indicator of vitamin D status (Holick et al., 2011). To measure serum 25(OH)D level, blood samples were taken after the participants had fasted for ≥ 8 hours and were assayed within 24 hours of transportation. Serum 25(OH)D levels were measured by radioimmunoassay (Diasorin, Stillwater, MN, USA) using a gamma counter (1470 Wizard Gamma Counter; Perkin Elmer, Turku, Finland). To minimize analytical variation, serum 25(OH)D levels were analyzed at the same institute, at which a quality assurance program was implemented throughout the analysis period. Serum 25(OH)D level was categorized according to quartiles (1.98-11.69, 11.70-15.0, 15.01-19.41, 19.42-60.47 ng/mL). Vitamin D deficiency was defined by serum 25(OH)D level below 20 ng/mL (Holick, 2007).

### 2.4. Potential covariates

Sociodemographic characteristics examined as covariates included age, education (<12 years or ≥12 years), marital status (married, living together, divorced/separated/widowed, or not married), and low monthly household income (≤ 25^th^ percentile of the national population). Lifestyle factors included current smoking, alcohol consumption, and physical activity. The Alcohol Use Disorder Identification Test Alcohol Consumption (AUDIT-C) instrument (Gordon et al., 2001) was used to assess severity of alcohol consumption, with a score of ≥8 indicating significant alcohol use problems (Seong et al., 2009). Physical activity was determined according to metabolic equivalent of task (MET) values, based on the self-reported frequency and duration of vigorous activity, moderate activity, and walking during the previous week. The MET value of a particular activity (vigorous activity = 8.0 MET; moderate activity = 4.0 MET; walking = 3.3 MET) was multiplied by the mean time (hours/week) spent performing that particular activity to calculate the MET-hours per week. The total weekly physical activity level was the sum of the weekly MET-hours for each activity (Ainsworth et al., 2000). Body weight status was determined by body mass index (BMI) and was categorized as <18.5, 18.5 to <23, 23 to <25, and ≥25 kg/m^2^, which are the cut-off points for underweight, normal, overweight, and obese for Asian populations (Kanazawa et al., 2005). The presence of chronic illness was assessed by self-report of a clinical diagnosis made by a physician and included the following diseases: diabetes mellitus, stroke, ischemic heart disease, renal failure, chronic hepatitis, and cancer. Serum creatinine was measured using the colorimetric method (Automatic Analyzer 7600; Hitachi, Tokyo, Japan), and the glomerular filtration rate (GFR) was then calculated using the Modification of Diet in Renal Disease (MDRD) method (Levey et al., 1999). The participants were defined as having past history of depression if they reported being clinically diagnosed with depression by a physician at some point during their lives.

### 2.5. Statistical analysis

Statistical analysis was conducted using Stata version 16.0 (StataCorp, College Station, TX, USA). Proportions and means were estimated for the demographic characteristics of the study sample. Logistic regression was used to estimate unadjusted and adjusted odds ratios (OR) and 95% confidence intervals (95% CI). Statistical significance was based on an α <0.05. As the KNHANES data were acquired using a complex sample, analyses incorporated an indicator for sample stratification, sampling weights and PSUs in all analyses except for the quantile regression where this was not possible. These analyses were implemented using the survey (“svy” commands in Stata v16, College Station, Tx).

The vitamin D status was defined by categorizing the continuous 25(OH)D level into quartiles. Logistic regression was used to assess the association of the lowest quartile concentrations of serum 25(OH)D status with the presence of depression as defined by PHQ-9 score ≥10. Quantile regression (75^th^ percentile) was used to assess the association using the continuous PHQ-9 score.

Models adjusted for age, sex, marital status, level of education, lowest income quartile, BMI, level of physical activity, chronic conditions, serum creatinine level, glomerular filtration rate (GFR), and past history of depression. Using forward selection, potential modification was assessed by including interaction terms in the model. Effect modification was considered to be present if the Wald Test was statistically significant (p <0.05). In the absence of effect modification, confounding was assessed. Adjustments leading to meaningful changes in the estimated regression coefficient for the association were interpreted as indicative of confounding.

## 3. Results

### 3.1. General characteristics of study population

Figure 1 shows the flow diagram for participants in this study. Of the 9,701 targeted individuals in KNHANES 2014, 7,550 participated in the survey. Of the 5,976 participants aged ≥19 years, serum vitamin D levels were collected for 2,018. In addition, 193 participants with missing values for PHQ-9 were excluded, leaving a total of 1,825 participants (859 men and 966 women) for cross-sectional analysis.

**Figure 1.**
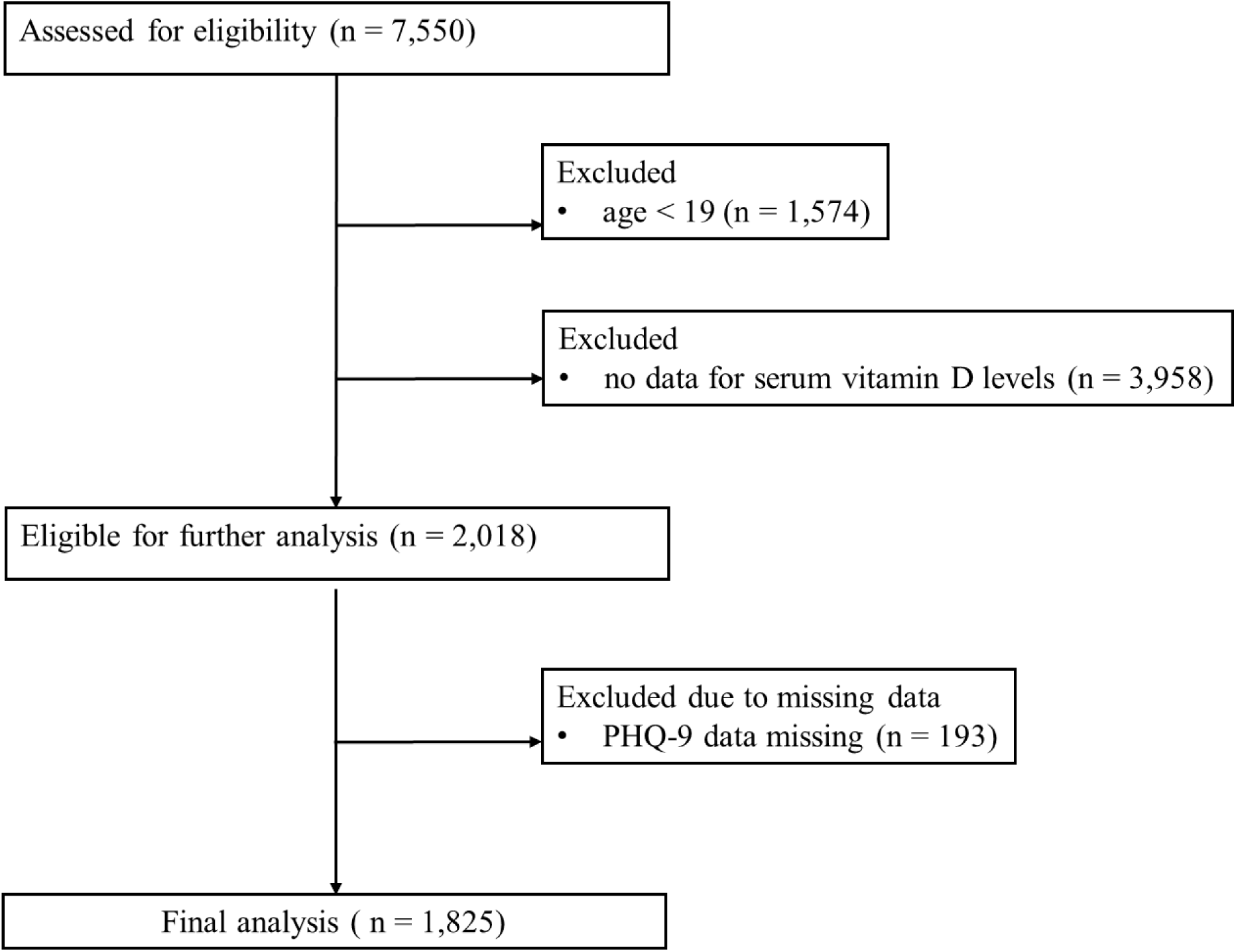
Flow diagram of the numbers of participants enrolled.

Table 1 presents the general characteristics of the study participants with and without depression. In this sample, a total of 116 cases of depression were identified, with an unweighted prevalence (95% CI) estimated as 6.4% (5.3%, 7.6%) in the unweighted data. Sensitivity analysis comparing unweighted and weighted estimates for various variables found the estimates to be similar. For example, the prevalence of depression, accounting for design effects, was 6.0%, compared to 6.4% in the unweighted data. Similarly, weighted mean 25(OH)D level was estimated as 15.8 ng/mL, compared to mean estimates of 16.2 ng/mL in the unweighted data.

**Table 1.**
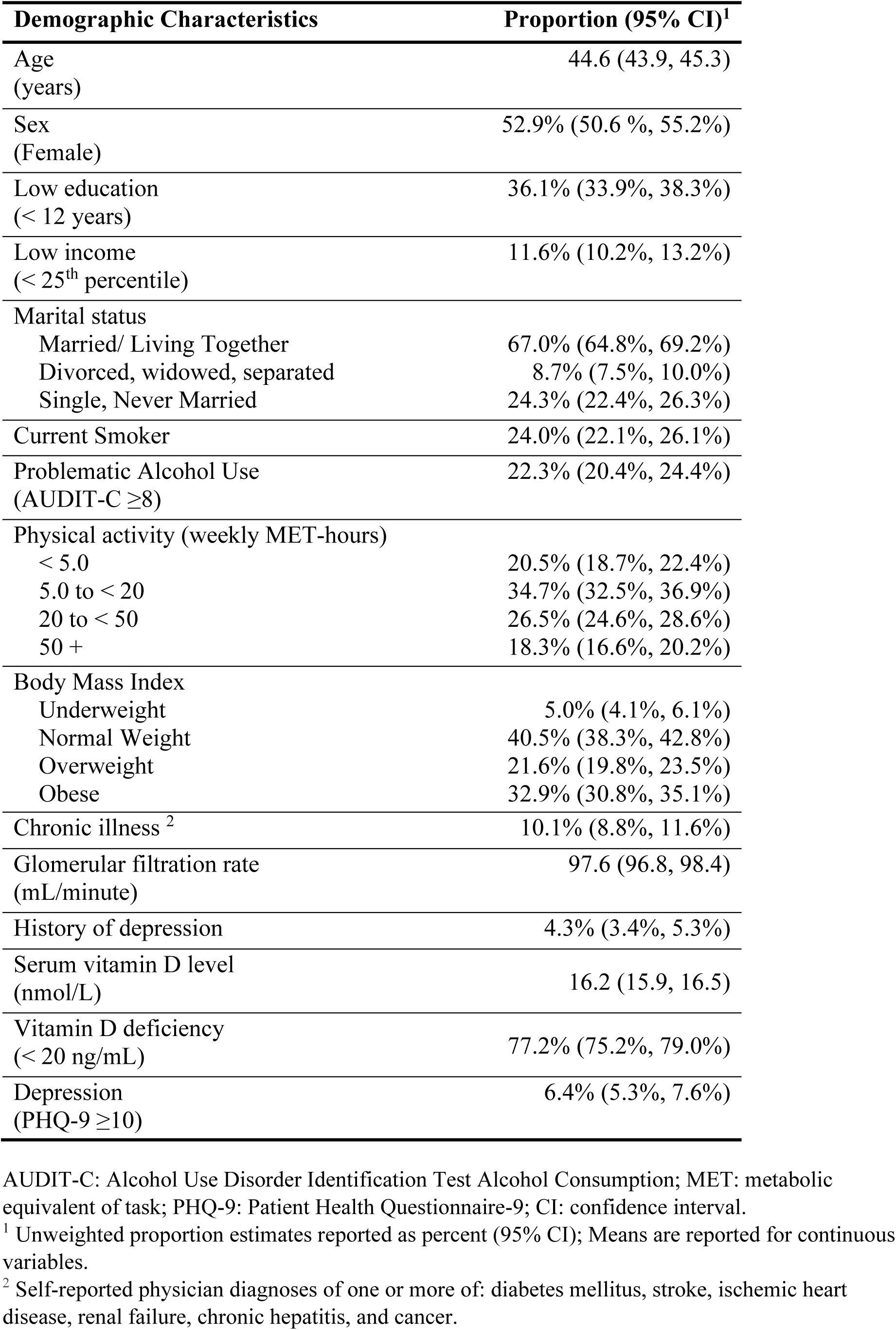
Estimates of the unweighted proportions of the demographic characteristics of the sample population.

| **Demographic Characteristics** | **Proportion (95% CI)<sup>1</sup>** |
| --- | --- |
| Age (years) | 44.6 (43.9, 45.3) |
| Sex (Female) | 52.9% (50.6 %, 55.2%) |
| Low education (< 12 years) | 36.1% (33.9%, 38.3%) |
| Low income (< 25<sup>th</sup> percentile) | 11.6% (10.2%, 13.2%) |
| Marital status |  |
| Married/ Living Together | 67.0% (64.8%, 69.2%) |
| Divorced, widowed, separated | 8.7% (7.5%, 10.0%) |
| Single, Never Married | 24.3% (22.4%, 26.3%) |
| Current Smoker | 24.0% (22.1%, 26.1%) |
| Problematic Alcohol Use (AUDIT-C ≥8) | 22.3% (20.4%, 24.4%) |
| Physical activity (weekly MET-hours) |  |
| < 5.0 | 20.5% (18.7%, 22.4%) |
| 5.0 to < 20 | 34.7% (32.5%, 36.9%) |
| 20 to < 50 | 26.5% (24.6%, 28.6%) |
| 50 + | 18.3% (16.6%, 20.2%) |
| Body Mass Index |  |
| Underweight | 5.0% (4.1%, 6.1%) |
| Normal Weight | 40.5% (38.3%, 42.8%) |
| Overweight | 21.6% (19.8%, 23.5%) |
| Obese | 32.9% (30.8%, 35.1%) |
| Chronic illness <sup>2</sup> | 10.1% (8.8%, 11.6%) |
| Glomerular filtration rate (mL/minute) | 97.6 (96.8, 98.4) |
| History of depression | 4.3% (3.4%, 5.3%) |
| Serum vitamin D level (nmol/L) | 16.2 (15.9, 16.5) |
| Vitamin D deficiency (< 20 ng/mL) | 77.2% (75.2%, 79.0%) |
| Depression (PHQ-9 ≥10) | 6.4% (5.3%, 7.6%) |

AUDIT-C: Alcohol Use Disorder Identification Test Alcohol Consumption; MET: metabolic equivalent of task; PHQ-9: Patient Health Questionnaire-9; CI: confidence interval.
<sup>1</sup> Unweighted proportion estimates reported as percent (95% CI); Means are reported for continuous variables.
<sup>2</sup> Self-reported physician diagnoses of one or more of: diabetes mellitus, stroke, ischemic heart disease, renal failure, chronic hepatitis, and cancer.

### 3.2. Association between serum 25(OH)D level and depression

The mean serum 25(OH)D level was higher in the non-depressed participants compared to those with depression (15.9 ng/mL versus 14.1 ng/mL), with a mean difference of −1.85 ng/mL (95% CI −3.06, −0.64; p =0.003). When analyzing quartiles of serum 25(OH)D, prevalence of depression in the lowest quartile was 8.3%, and in the upper three quartiles was 5.1%, with an unadjusted OR of 1.67 (95% CI 1.07, 2.62; p =0.024). Similarly, the vitamin D deficient group had a higher estimated prevalence of depression compared to the non-deficient group (6.3% versus 4.6%), although this difference was not statistically significant (OR = 1.40, 95% CI 0.82, 2.40; p =0.216). The remaining analyses used the vitamin D exposure dichotomized by the lowest concentration quartile.

### 3.3. Potential modification

One interaction was identified in the logistic regression analysis, a sex by vitamin D exposure interaction. When the models included a female sex interaction term, the regression coefficient was −1.15, and in exponentiated form (as an OR) was 0.32 (p = 0.028). Upon stratification, the OR in men was 3.21 (95% CI 1.45, 7.12; p = 0.004) and in women the OR was 1.02 (95% CI 0.57, 1.80; p = 0.957). In view of the large number of interactions evaluated, the lack of a biological explanation for such a sex difference, and the lack of an interaction in the quantile regression analysis (see below), we interpreted the significance of this interaction term as a probable Type I error and conducted the multivariable analysis, described below, in the combined sample of both men and women. Notably, the interaction was not observed in a logistic regression model using the criteria for vitamin D deficiency (below 20 ng/mL) rather than the lowest quartile (OR = 1.01, 95% CI 0.96, 1.06; p =0.71).

### 3.4. Potential confounding

Potential confounding was assessed using forward selection. The results are presented in Table 2. Potential covariates of interest included sex, marital status, alcohol use (which weakens the observed association) and history of depression (which strengthens the association). In a model simultaneously adjusting for these variables, the OR for serum 25(OH)D diminished to 1.21 (95% CI 0.74-1.99, p = 0.448). In this multivariable model, problematic alcohol use was no longer associated with depression (adjusted OR for problematic alcohol use = 1.53, 95% CI 0.84, 2.80; p = 0.166). With removal of this variable the OR for serum 25(OH)D was 1.48 (95% CI 0.93, 2.35; p = 0.097). The model is presented in Table 3. Notably, when the analysis is restricted to male respondents, the association remained significant with these adjustments: the OR for serum 25(OH)D adjusted for marital status, problematic alcohol use and past history of depression remained significant (OR = 2.54, 95% CI 1.17, 5.50; p = 0.018).

**Table 2.** Estimates of the unadjusted and adjusted logistic regression models.

| **Demographic Characteristics** | **Unadjusted OR**<br>**(95% CI)** | **Adjusted OR**<br>**(95% CI)** |
| --- | --- | --- |
| Age<br>(years) | 0.99 (0.98, 1.00) | 1.64 (1.05, 2.56) |
| Sex<br>(Female) | 2.26 (1.35, 3.78) | 1.50 (0.94, 2.40) |
| Low education<br>(< 12 years) | 0.80 (0.51, 1.27) | 1.65 (1.04, 2.60) |
| Low income<br>(< 25<sup>th</sup> percentile) | 4.14 (2.51, 6.48) | 1.68 (1.07, 2.63) |
| Marital status |  |  |
| Married/ Living Together | Reference | 1.48 (0.97, 2.28) |
| Divorced, widowed, separated | 2.89 (1.59, 5.26) |  |
| Single/ Never Married | 1.73 (1.13, 2.66) |  |
| Current Smoker | 1.23 (0.77, 1.97) | 1.67 (1.07-2.62) |
| Problematic Alcohol Use<br>(AUDIT-C ≥ 8) | 1.12 (0.65, 1.93) | 1.45 (0.88, 2.38) |
| Physical activity (weekly MET-hours) |  |  |
| < 5.0 | 1.12 (0.59, 2.60) | 1.64 (1.05, 2.60) |
| 5.0 to < 20 | 1.02 (0.58, 1.79) |  |
| 20 to < 50 | 0.72 (0.37, 1.39) |  |
| 50 + | Reference |  |
| Body mass index |  |  |
| Underweight | 3.34 (1.59, 7.03) | 1.62 (1.03, 2.55) |
| Normal Weight | Reference |  |
| Overweight | 1.36 (0.67, 2.77) |  |
| Obese | 1.27 (0.75, 2.18) |  |
| Chronic Illness * | 1.92 (1.02, 3.63) | 1.67 (1.07, 2.59) |
| Glomerular Filtration Rate<br>(mL/minute) | 1.00 (0.99, 1.01) | 1.68 (1.07, 2.65) |
| Past history of depression | 11.5 (6.4, 20.6) | 1.86 (1.17, 2.96) |

AUDIT-C: Alcohol Use Disorder Identification Test Alcohol Consumption; MET: metabolic equivalent of task; OR: odds ratios; CI: confidence interval.
\* Self-reported physician diagnoses of at least one of: diabetes mellitus, stroke, ischemic heart disease, renal failure, chronic hepatitis, and cancer.

**Table 3.** Logistic regression model including adjusting for sex, marital status, and history of depression.

|  | **OR (95% CI)** | ***p*-value** |
| --- | --- | --- |
| 25(OH)D: lowest quartile | 1.48 (0.93, 2.35) | 0.097 |
| Sex (female) | 2.06 (1.21, 3.51) | 0.008 |
| Married/ Living Together | Reference |  |
| Divorced, Widowed, Separated | 2.08 (1.12, 3.87) | 0.021 |
| Single/Never married | 1.86 (1.19, 2.89) | 0.007 |
| Past-history of depression | 9.59 (5.24, 17.56) | <0.001 |

\* Estimates presented as Odds Ratios (OR) and 95% CI (95% CI).

### 3.5. Quantile regression analysis

Although adjustment for the complex sampling strategy was not possible using this technique, quantile regression was used to evaluate the association using the continuous PHQ-9 scores. The 75^th^ percentile was used as the index quantile in this analysis. The unadjusted effect of serum 25(OH)D was statistically significant (p = 0.007) and remained significant after individual adjustments for each of the covariates listed in Table 2. The sex by serum 25(OH)D interaction seen in the logistic regression analysis was not significant (β = 0.059, p = 0.182). With simultaneous adjustment for sex, marital status, alcohol use and past history of depression, the effect of serum 25(OH)D remained statistically significant (β = −0.056, p = 0.002).

## 4. Discussion

These analyses demonstrate an association between low serum vitamin D levels and moderate/severe depressive symptoms, in a nationally representative sample of the adult Korean population. Additionally, this association appears to be stronger among males than females.

Our results are consistent with those of previous cross-sectional studies showing an inverse association between serum 25(OH)D levels and depressive symptoms (Chan et al., 2011; Ganji et al., 2010; Hoang et al., 2011; Hoogendijk et al., 2008; Jaaskelainen et al., 2015; Jovanova et al., 2017; Kjaergaard et al., 2011; Song et al., 2016; Stewart and Hirani, 2010). Although many of these studies reported a positive association between vitamin D deficiency and depression in older adults (Chan et al., 2011; Hoogendijk et al., 2008; Jovanova et al., 2017; Song et al., 2016; Stewart and Hirani, 2010), there is less empirical support for this association in adult samples of all ages. A study of a Norwegian population (10,086 adults aged 30–87 years) found that low serum 25(OH)D levels were a significant predictor of depressive symptoms in both smokers and non-smokers (Kjaergaard et al., 2011). Similar results were also reported by a study of a sample of 12,594 residents (aged 20–95 years) in the United States. This study found that higher serum 25(OH)D levels were associated with a significantly decreased risk of current depressive symptoms (Hoang et al., 2011). According to the Health 2000 Survey of Finland, higher serum 25(OH)D levels were associated with a lower prevalence of depressive disorders in 5,371 adults aged 30–79 years (Jaaskelainen et al., 2015).

However, these results are not conclusive as there have also been several findings of no association between vitamin D status and depression after adjusting for various confounding factors (Nanri et al., 2009; Pan et al., 2009; Park et al., 2016; Zhao et al., 2010). Zhao et al. (2010) conducted a large cross-sectional, population-based study of adults aged ≥20 years in the United States (n=3916), which found no association between serum 25(OH)D and depression (PHQ-9 ≥10) after adjustment for confounding factors (Zhao et al., 2010). In the fifth KNHANES, serum 25(OH)D levels were not associated with depression in Korean adults (Park et al., 2016). However, a major limitation was that depression was assessed by a single question in that study: “Have you felt sad or hopeless for at least 2 consecutive weeks during the past year to the extent that you had difficulty performing your usual activities?”

Our present findings show that vitamin D deficiency in Korea is prevalent and seems to be higher than in western studies (Holick, 2017). A total of 77.2% of our sample met criteria for vitamin D deficiency, serum 25(OH)D level <20 ng/mL, which is the minimal level of serum 25(OH)D that minimizes the risk for rickets (Holick, 2007). Vitamin D deficiency is now well recognized as a more common health problem in Asian countries. A variety of cultural, environmental, and genetic factors such as darker skin, differences in diet and less use of supplemental vitamin D have been suggested as factors influencing vitamin D status in Asian countries compared to western countries (Park et al., 2018). Vitamin D is a unique vitamin in that it is mostly acquired by cutaneous synthesis in response to sunlight exposure, as dietary sources of vitamin D are minimal (Holick, 2017). Given the association between low vitamin D status and depression, interventions to increase vitamin D levels, such as supplementation and increasing exposure to sunlight, should be considered for depressed individuals at risk for vitamin D deficiency.

This analysis found evidence of an association between vitamin D status and depression. However, the result should be interpreted with caution for two reasons. First, a sex by serum 25(OH)D interaction term was significant in some parts of the analysis. The interaction suggested a stronger effect of serum 25(OH)D in males than females, with stratified analysis finding a significant effect in males and an OR near the null value in females. In the absence of a biological explanation for this, and in view of the multiple interactions examined, and due to a lack of consistency with other parts of the analysis, we interpreted this as a probable type 1 error rather than an indicator of effect modification. However, the possibility of effect modification by sex should be kept in mind by future investigators studying this question.

A few previous studies also reported that the association between serum 25(OH)D and depressive symptoms was stronger in males than in females. In a representative sample of the Finnish adult population, higher serum 25(OH)D levels were associated with a lower prevalence of depressive disorders, especially among younger men (Jaaskelainen et al., 2015). In an older Korean population, serum 25(OH)D levels were inversely associated with depressive symptoms in men but not in women (Song et al., 2016). Additionally, a large Australian epidemiologic study of young adults reported that each 10 nmol/L increase in serum 25(OH)D levels was associated with a decrease of 8% in depression scores in males (but not in females) (Black et al., 2014). Sex-specific confounders, such as sex hormones, may account for differences in the effect of low serum 25(OH)D in males and females. Vitamin D is known to affect androgen synthesis in testicular cells in males (Hofer et al., 2014), and higher testosterone levels have protective benefits against depression (McHenry et al., 2014). In this context, the concurrent presence of low vitamin D and low testosterone levels may be synergistically associated with depressive symptoms among males. A previous study reported a stronger immunomodulatory effect of vitamin D in females than in males, suggesting a synergy between vitamin D and estradiol (Correale et al., 2010).

Second, adjustment for confounding variables weakened the observed association, providing evidence of possible confounding. The association of vitamin D and depression was not statistically significant after adjustment for sex, marital status and problematic alcohol use in the logistic regression analysis. While this was not seen in the quantile regression analysis, it indicates that additional research is needed to confirm that the effect of serum 25(OH)D on depression is independent of these variables. Also, as these are cross-sectional associations, they cannot support causal assertions. In terms of confounding variables, several previous studies have reported that lower serum 25(OH)D levels were associated with depression, particularly among those with low socioeconomic status or unhealthy lifestyle (e.g., problematic alcohol use) (Jaaskelainen et al., 2015; Song et al., 2016), which is partially consistent with our findings. This may suggest that vitamin D provides protection against the development of depression. Conversely, low vitamin D status may also be the consequence of depression, as the impact of depression on health-related factors, such as physical activity, alcohol use, smoking, diet, and obesity, have been linked to vitamin D status (Lotfaliany et al., 2019; Luppino et al., 2010; van Dam et al., 2007). More evidence from prospective or intervention studies regarding vitamin D may be needed to elucidate any causal relationships and clarify the effects of potential confounding factors.

The strengths of our study include the large, nationally representative sample and the use of statistical procedures that were designed to produce accurate estimates from population-based survey data. In addition, the availability of a wide range of information in KHANES allowed us to clarify the association between serum 25(OH)D levels and depression after adjustment for various covariates.

The present study also has several limitations. First, as noted herein, the cross-sectional nature of our design limited our interpretation of the results, as this design precluded determinations of the direction of potential causal associations. Another limitation was the use of the PHQ-9, a self-rated depressive symptom scale, to categorize depression instead of using a standardized diagnostic interview. Although the PHQ-9 has been well-validated as a screening tool for depression, it is not a diagnostic tool. Although our findings may not precisely reflect the population of individuals with depression who also have low vitamin D status, our results show that individuals with elevated depressive symptoms also have low serum 25(OH)D levels. Finally, regular dietary or supplementary intake of vitamin D, as well as sunlight exposure, time spent indoors, seasonal variations, the use of medications, and other confounding variables known to be related to vitamin D and depression (Holick, 2007), were not considered. Moreover, data that pose a risk of disclosure of the identity of a participant, such as the timing of data collection, residential area, and occupations were not available in the KNHANES.

## 5. Conclusion

This study found a cross-sectional association between low serum 25(OH)D levels and depressive symptoms in a community-living Korean adult population, which was more prominent in males than in females. Further studies are needed to examine the relationship between vitamin D and depression, to explore sex differences in this regard, and to investigate the biological and behavioral mechanisms involved.

## Data Availability

Data came from a publicly available dataset as per the manuscript details.

## CRediT authorship contribution statement

**Young-Eun Jung:** Conceptualization, Writing - original draft. **Ashley K. Dores:** Formal analysis, Writing - original draft. **Scott B. Patten:** Formal analysis, Writing – Review & Editing. **Lakshmi N. Yatham:** Writing – Review & Editing. **Raymond W. Lam:** Conceptualization, Writing – Review & Editing.

